# Homocysteine thiolactone contributes to a prognostic value of fibrin clot structure/function in coronary artery disease patients

**DOI:** 10.1101/2022.07.12.22277430

**Authors:** Marta Sikora, Paweł Skrzydlewski, Joanna Perła-Kaján, Hieronim Jakubowski

## Abstract

**Background:** Fibrin clot structure/function contributes to cardiovascular disease. We examined sulfur-containing metabolites as determinants of fibrin clot lysis time (CLT) and maximum absorbance (Abs_max_) in relation to outcomes in coronary artery disease (CAD) patients. Effects of B-vitamin/folate therapy on CLT and Abs_max_ were studied.

**Methods and Findings:** Plasma samples were collected from 1,952 CAD patients randomized in a 2 × 2 factorial design to (*i*) folic acid, vitamins B_12_, B_6_; (*ii*) folic acid, vitamin B_12_; (*iii*) vitamin B_6_; (*iv*) placebo for 3.8 years in the Western Norway B-Vitamin Intervention Trial. Clot lysis time (CLT) and maximum absorbance (Abs_max_) were determined using a validated turbidimetric assay. Acute myocardial infarction (AMI) and mortality were assessed during a 7-year follow-up. Data were analyzed using bivariate and multiple regression. Survival free of events was studied using Kaplan Mayer plots. Hazard ratios (HR) and 95% confidence intervals (CI) were estimated using Cox proportional hazards models. Baseline urinary (u)Hcy-thiolactone and plasma Cys were significantly associated with CLT while plasma Hcy was significantly associated with Abs_max_, independently of fibrinogen, triglycerides, vitamin E, glomerular filtration rate, body mass index, age, sex plasma creatinine, CRP, HDL-C, ApoA1. B-vitamins/folate did not affect CLT and Absmax. Kaplan-Meier analysis showed associations of increased baseline CLT and Abs_max_ with worse outcomes. In Cox regression analysis, baseline CLT and Abs_max_ (>cutoff) predicted AMI (CLT: HR 1.58, 95% CI 1.10-2.28; *P* = 0.013. Abs_max_: HR 3.22, CI 1.19-8.69; *P* = 0.021) and mortality (CLT: HR 2.54, 95% CI 1.40-4.63; *P* = 0.002. Abs_max_: 2.39, 95% CI 1.17-4.92; *P* = 0.017). After adjustments for other prognostic biomarkers these associations remained significant.

**Conclusions:** uHcy-thiolactone and plasma Cys are novel determinants of CLT, an important predictor of adverse CAD outcomes. CLT and Abs_max_ were not affected by B-vitamin/folate therapy, which could account for the lack of efficacy of such therapy in CAD.

**Trial Registration:** URL: http://clinicaltrials.gov. Identifier: NCT00354081

## Introduction

Thrombotic events initiated by an underlying vascular dysfunction are a major component of cardiovascular disease (CVD). In occlusive arterial disease the formation of a platelet-rich thrombus is supported by a fibrin mesh, whose formation depends on complex interactions between the components of the coagulation cascade. Accumulating evidence suggests that fibrin clot structure and function is associated with the development and progression of CVD. CAD patients. For example, dense structure of the fibrin clot, reflected in increased maximum absorbance and longer clot lysis time, have been observed in CVD patients [1]. Fibrin clot formation and lysis are dynamic processes and identification of factors affecting complex phenotypes reflecting fibrin structure and function is important in assessing CVD risk and the development of new treatments [2].

Elevated plasma total homocysteine (tHcy), *i*.*e*. hyperhomocysteinemia (HHcy), increases a risk for the development of CVD and stroke [3, 4]. tHcy is a composite marker that includes mostly disulfides such as *S*-Hcy-albumin, *S*-Hcy-IgG, and Hcy-S-S-Cys, with free reduced Hcy contributing only about 1% [3, 4]. However, it should be noted that other Hcy metabolites, such as Hcy-thiolactone and *N*-Hcy-protein [5], which have been independently implicated in CVD [6] and stroke [7], are not accounted for by the tHcy marker [3].

The only known source of Hcy, an important intermediate in one-carbon metabolism in humans, is the dietary protein methionine. Hcy is metabolized to Hcy-thiolactone in a reaction catalyzed by methionyl-tRNA synthetase. Hcy-thiolactone, a chemically reactive thioester, modifies protein lysine residues generating *N*-Hcy-protein [5]. Hcy-thiolactone concentrations in healthy human subjects are about 100-fold higher in urine (median 144 nM, range 11 - 485 nM) [8] than in plasma (median 0.56 nM, range <0.1 - 22.6 nM) [9]. Urinary (u)Hcy-thiolactone can be as high as 2-4 μM in CAD patients [6], and 10-15 μM in severely HHcy *Cbs*^-/-^ mice [10]. A prospective, randomized clinical intervention study showed that uHcy-thiolactone predicted acute myocardial infarction (AMI) in a cohort of coronary artery disease (CAD) patients and that therapy with any combination of folic acid, B_12_, and B_6_ vitamins did not affect levels of uHcy-thiolactone and its association with AMI [6].

How sulfur-containing metabolites affect fibrin clot structure/function has not been examined in large randomized clinical trials. For this reason, we quantified fibrin clot lysis time (CLT, a measure of clot function) and maximum absorbance (Abs_max_, a measure of clot structure) in a cohort of CAD patients participating in the Western Norway B-Vitamin Intervention Trial (WENBIT). We have studied associations of fibrin CTL and Abs_max_ with Hcy-thiolactone, Hcy and cysteine, and effects of folic acid and B-vitamin therapy on these variables. We also examined how these associations are influenced by other variables related to CVD risk and examined prognostic values of CLT and Abs_max_.

## Methods

### Patients

We analyzed existing citrated plasma samples from patients with suspected CAD who underwent coronary angiography for stable angina pectoris and participated in WENBIT [11]. Participant characteristics and blood/urine samples, collected at baseline and median 38-month follow-up, have been previously described [11]. Briefly, most participants (90%) had significant coronary stenosis (>50% of cross-surface area obstructed in at least one of the major coronary arteries), cardiovascular history/risk factors (60 %), and were on medications during the trial, including antiplatelet drugs (92%), acetyl salicylic acid (90.2), statins (88.4%), and β-blockers (78.2), following baseline angiography. Participants were randomly assigned to groups receiving (*i*) folic acid (0.8 mg) + vitamin B_12_ (cyanocobalamin, 0.4 mg), vitamin B_6_ (pyridoxine, 40 mg); (*ii*) folic acid + vitamin B_12_; (*iii*) vitamin B_6_; (*iv*) placebo for an average of 3.8 years. The study medication (Alpharma Inc, Copenhagen, Denmark) was given as a single capsule, indistinguishable by color, weight, or the ability to dissolve in water.

The present study included 61.2-year-old patients (71.5% male) from baseline (n = 1,952) and the end-of-study (n = 192) for whom uHcy-thiolactone values were available. Samples were assayed by investigators blinded to the clinical data to avoid bias. The study protocol was approved by the Regional Committee for Medical and Health Research Ethics, Bergen, Norway; by the Norwegian Medicines Agency, Bergen, Norway; and the WENBIT Steering Committee, Bergen, Norway.

### Clinical endpoints

The endpoints were mortality and AMI, which included both fatal and nonfatal events, defined according to the International Classification on Diseases (ICD) 10th edition; I21-22. Information on endpoints was obtained from the Cardiovascular Disease in Norway (CVDNOR; https://cvdnor.b.uib.no/) project, which provided information on discharge diagnoses from Norwegian hospitals during 1994-2009, linked to each patient’s unique 11-digit personal number [12].

### The clotting/lysis assay

The assay was modified from that described previously [2]. Briefly, 25 µL citrated plasma was added to 75 µL buffer (50 mM Tris-HCl, 150 mM NaCl, pH 7.6) containing 12.5 ng of tPA (Molecular Innovations), 83 ng/ml final concentration. The reaction was initiated with 50 µl of activation mix containing 0.09 U/mL of thrombin (Millipore-Sigma) and 22.5 mM CaCl_2_ in 50 mM Tris-HCl, 150 mM NaCl (pH 7.6) to each well of the 96-well plate using a multichannel pipette at 20 sec intervals. The time of addition of the activation mix was recorded to enable the plate reader times to be adjusted to the start of clot initiation. Absorbance was read at 340 nm every 30 s for 1 hour in a Tecan NanoQuant Infinite M200 Pro microplate reader. Complete lysis of fibrin clots occurred within 1 hour at the tPA concentration used. Each plasma sample was assayed in duplicate.

### Clotting/Lysis Data Analysis

Kinetics of fibrin clot formation and lysis, illustrated in **Figure S1**, were analyzed using a customized software kindly provided by Dr. Peter Grant [2]. Maximum absorbance at 340 nm (Abs_max_, a measure of fibrin network density) and fibrin CLT (*i*.*e*., a time it took for Abs_max_ to drop by 50%, a measure clot’s susceptibility to lysis) were calculated from the kinetics. Inter-assay variabilities for Abs_max_ and fibrin CLT were 1.5% and 4.7%, respectively. The definitions of clotting and lysis variables are shown in **Figure S1**. Correlations between the clotting and lysis variables in the WENBIT cohort, shown in **Table S1**, are like the correlations previously reported in healthy individuals [2].

### Metabolite and other variable assays

Values for serum/plasma tHcy, Cys, creatinine, folate, vitamins B_6_ and B_12_, urinary Hcy-thiolactone and creatinine, and other variables were obtained from analyses reported previously [6, 11].

### Statistics

Normality of distribution was tested with the Shapiro-Wilk’s statistic. Mean ± standard deviation (SD) or median was calculated for normally or non-normally distributed variables, respectively. An unpaired two-sided *t*-test was used for comparisons between two groups of variables with normal distribution. A Mann-Whitney rank sum test was used for comparisons between two groups of non-normally distributed variables. Associations between fibrin clot lysis time (CLT) or Abs_max_ and other variables were studied by bivariate and multiple regression analyses. Receiver-operating characteristic (ROC) curves were created to assess the optimal cut-off values for CLT and Abs_max_. Event-free survival was analyzed by the Kaplan-Meier method and log-rank test was used to estimate differences in survival between patients with long vs. short fibrin CLT. Hazard ratios (HR) and 95% confidence intervals (CI) for clinical events associated with long vs. short fibrin CLT were calculated using the multivariable Cox proportional hazard regression analysis. Statistical software packages Statistica, version 13 (TIBCO Software Inc., Palo Alto, CA, USA) and PSPP, version 1.0.1 (www.gnu.org) were used. Probability values were 2-sided and *P* value <0.05 was considered significant.

## Results

### Baseline values of fibrin CLT and Abs_max_

For the CAD patients (n = 1,952) mean age at baseline was 61.2 years and 29% were women. Baseline fibrin CLT varied from 105 to 1560 s and was significantly longer in women (n = 556) than in men (n = 1396), *P* = 0.0097 (**Table 1**). Longer fibrin CLT in women was accompanied by reduced levels of uHcy-thiolactone, urinary creatinine (uCreatinine), plasma creatinine (pCreatinine), plasma tHcy (but not cysteine, Cys), and older age compared to men. Fibrin clot Abs_max_ varied from 0.0025 to 0.362 A_340_ and showed a tendency for lower values in women compared to men (*P* = 0.055, **Table 1**).

**Table 1.**
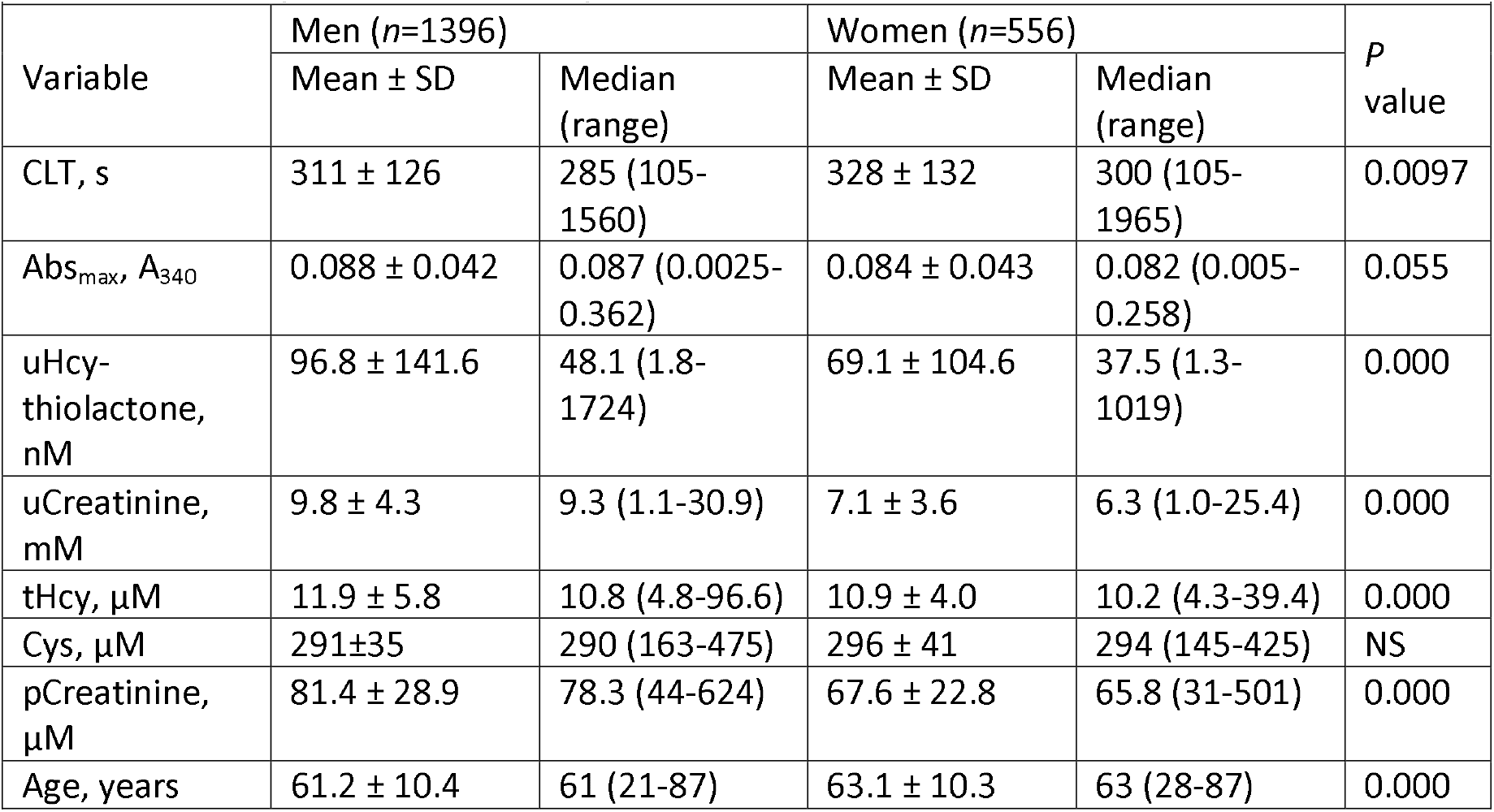
Baseline plasma fibrin clot lysis tome (CLT), maximum absorbance (Abs_max_), and other variables in CAD patients stratified by sex

| **Table 1** Baseline plasma fibrin clot lysis time (CLT), maximum absorbance (Abs<sub>max</sub>), and other variables in CAD patients stratified by sex |  |  |  |  |  |
| --- | --- | --- | --- | --- | --- |
| Variable | Men (n=1396) |  | Women (n=556) |  | p value |
|  | Mean ± SD | Median (range) | Mean ± SD | Median (range) |  |
| CLT, s | 311 ± 126 | 285 (105-1560) | 328 ± 132 | 300 (105-1965) | 0.0097 |
| Abs<sub>max</sub>, A<sub>340</sub> | 0.088 ± 0.042 | 0.087 (0.0025-0.362) | 0.084 ± 0.043 | 0.082 (0.005-0.258) | 0.055 |
| uHcy-thiolactone, nM | 96.8 ± 141.6 | 48.1 (1.8-1724) | 69.1 ± 104.6 | 37.5 (1.3-1019) | 0.000 |
| uCreatinine, mM | 9.8 ± 4.3 | 9.3 (1.1-30.9) | 7.1 ± 3.6 | 6.3 (1.0-25.4) | 0.000 |
| tHcy, μM | 11.9 ± 5.8 | 10.8 (4.8-96.6) | 10.9 ± 4.0 | 10.2 (4.3-39.4) | 0.000 |
| Cys, μM | 291±35 | 290 (163-475) | 296 ± 41 | 294 (145-425) | NS |
| pCreatinine, μM | 81.4 ± 28.9 | 78.3 (44-624) | 67.6 ± 22.8 | 65.8 (31-501) | 0.000 |
| Age, years | 61.2 ± 10.4 | 61 (21-87) | 63.1 ± 10.3 | 63 (28-87) | 0.000 |

### uHcy-thiolactone, glomerular filtration rate, and plasma Cys, but not tHcy, are correlated with fibrin CLT but not with Abs_max_

Relationships between plasma fibrin CLT and uHcy-thiolactone, plasma Cys and tHcy are illustrated in **Figure 1**. Fibrin CLT was significantly negatively correlated with uHcy-thiolactone (**Figure 1A**), glomerular filtration rate (GFR) (**Figure 1B**) and positively with plasma Cys (**Figure 1C**) but was unaffected by tHcy levels (**Figure 1D**). There was no correlation between fibrin Abs_max_ and uHcy-thiolactone, GFR, plasma Cys or tHcy (not shown).

**Figure 1.**
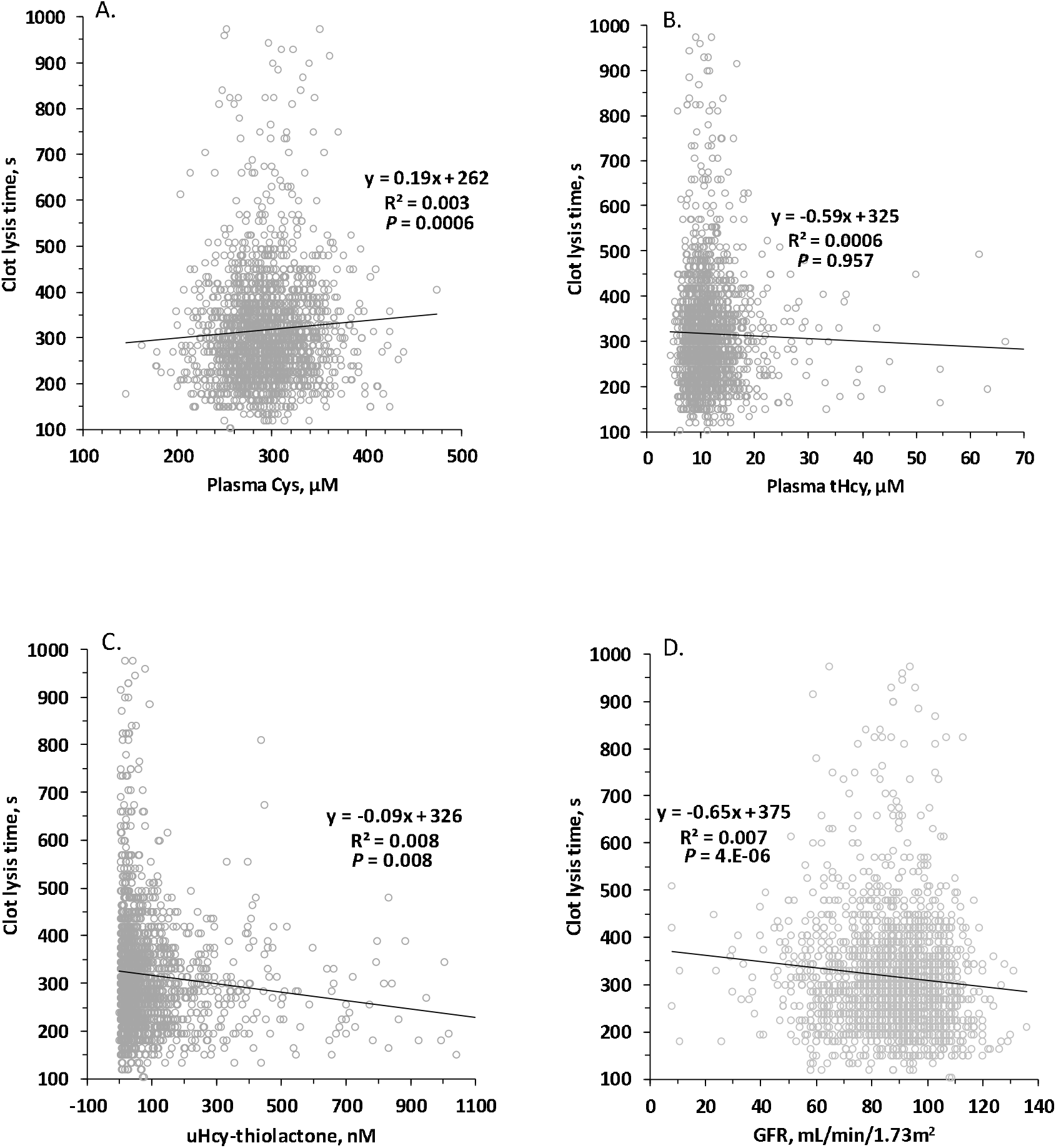
Relationships between CLT and plasma Cys (A.), tHcy (B.), uHcy-thiolactone (C.) and GFR (D.). Spearman *P* values are shown.

These findings suggest that better clearance of plasma Hcy-thiolactone into urine [8] shortens plasma fibrin CLT and thus is beneficial. On the other hand, higher plasma Cys appears to be detrimental because it prolongs plasma fibrin CLT.

### GFR affects plasma fibrin CLT as well as urinary Hcy-thiolactone and plasma Cys

These findings also suggest that impairments in kidney function and attenuation of GFR would reduce fibrinolysis. Indeed, we found that fibrin CLT was significantly negatively correlated with GFR: shorter at high GFR and longer at low GFR (**Figure 1B**). We also found that excretion of uHcy-thiolactone was better at high GFR and worse at low GFR (**Figure 2A**), mimicking better excretion of uCreatinine at high GFR and worse at low GFR (**Figure 2B**). Although we were not able to quantify plasma Hcy-thiolactone due to limited availability of samples, we found a negative correlation between plasma Cys and GFR (higher plasma Cys levels in CAD patients with low GFR, lower plasma Cys in high GFR patients; **Figure 2C**), similar to the correlation between pCreatinine and GFR (**Figure 2D**).

**Figure 2.**
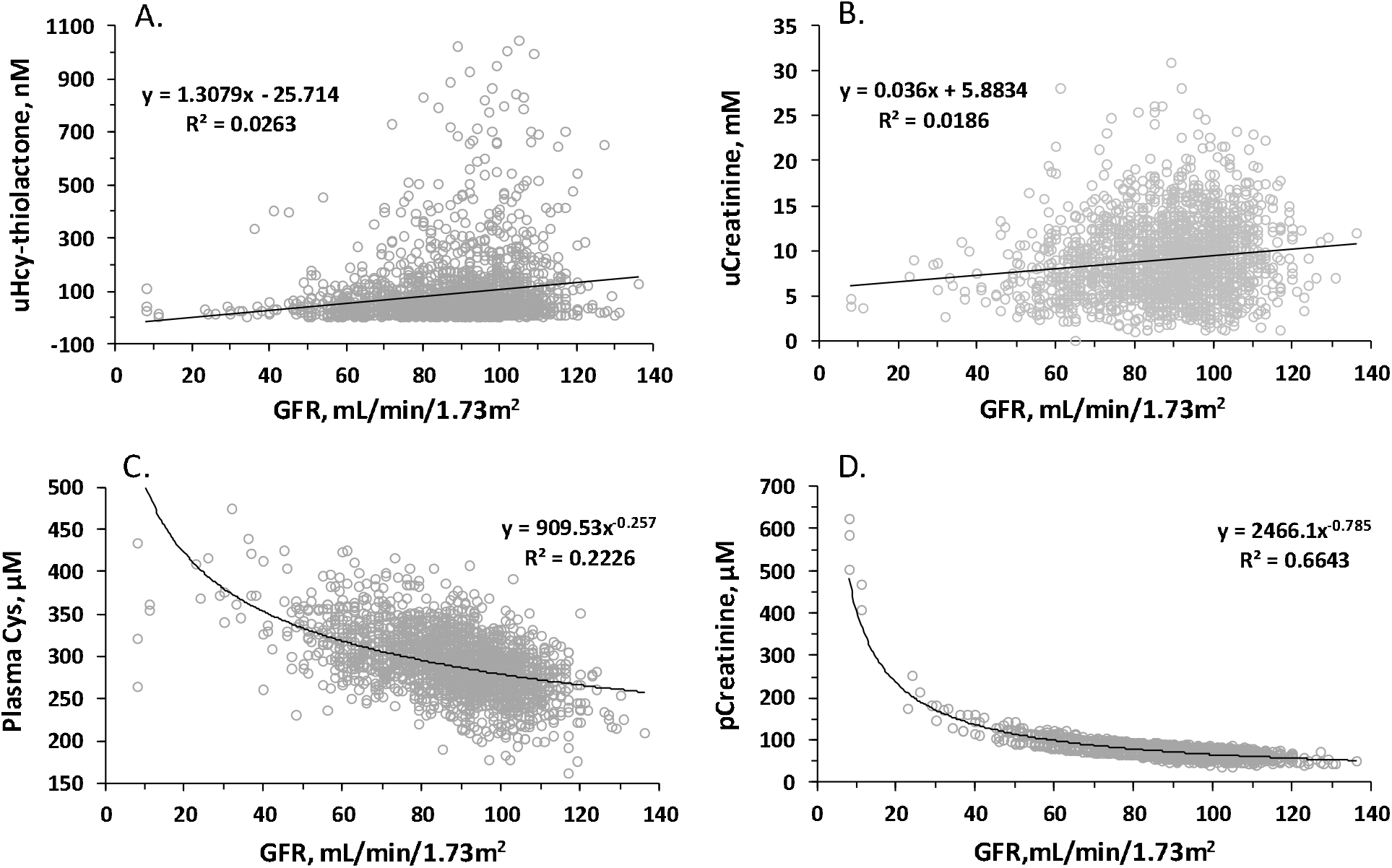
Relationships between GFR and uHcy-thiolactone (A.), uCreatinine (B.), plasma Cys (C.) and pCreatinine (D.).

### Other determinants of fibrin CLT and Abs_max_

ANOVA analysis showed the fibrin CLT and Abs_max_ were significantly correlated with each other, as previously shown in other studies [2][13][14]. Of the four examined sulfur-containing metabolites, Cys was positively associated with both fibrin CLT (**Table S1**) and Abs_max_ (**Table S2**). Hcy and methionine did not affect fibrin CLT or Abs_max_, while uHcy-thiolactone was negatively associated with fibrin CLT but not with Abs_max_ (**Table S1** and **S2**).

Both fibrin CLT (**Table S1**) and Abs_max_ (**Table S2**) were also associated with other variables: positively with fibrinogen, CRP, and age; negatively with GFR). In addition, fibrin CLT was associated with variables that did not affect Abs_max_: positively with vitamin E, triglycerides (TG), ApoB, Lpa, and BMI, suggesting that fibrinolysis was impaired by increases in these variables, and negatively with high density lipoprotein cholesterol (HDL-C) and uCreatinine. The negative association of fibrin CLT with HDL-C suggests that fibrinolysis was augmented by HDL-C. Conversely, Abs_max_ was associated with only one variable, pCreatinine, that did not affect fibrin CLT. Associations of fibrin CLT with fibrinogen, CRP, age, HDL-C, ApoB, Lpa, TG, and body mass index (BMI) have been previously reported by other investigators [1, 15][16][17][18][19].

Fibrin CLT and Abs_max_ showed disparate correlations with many other variables. For example, fibrin CLT significantly increased with increasing vitamin E (**Table S1**) while Abs_max_ was not affected by vitamin E (**Table S2**). Fibrin CLT significantly increased with increasing fibrinogen, C-reactive protein (CRP), TG, ApoB, and Lpa levels, as well as with BMI and age (**Table S1**), In contrast, fibrin CLT and Abs_max_ both significantly increased with decreasing GFR (**Table S1** and **S2**), indicating attenuated fibrinolysis in CAD patients with impaired kidney function. CLT significantly increased with decreasing uCreatinine, but not pCreatinine (**Table S1**). Abs_max_ increased with increasing pCreatinine but was unaffected by uCreatinine. In contrast to fibrin CLT, Abs_max_ was not affected by ApoB, Lpa, and age (**Table S2**).

Multiple regression analysis showed that fibrin CLT and Abs_max_ were significantly associated with each other. Of the four sulfur-containing metabolites, uHcy-thiolactone and Cys were negatively associated with fibrin CLT but not with Abs_max_ (**Table 2**). In contrast, Hcy was negatively associated with Abs_max_, but not with CLT (**Table 2**). Methionine did not affect fibrin CLT or Abs_max_ (not shown).

**Table 2.**
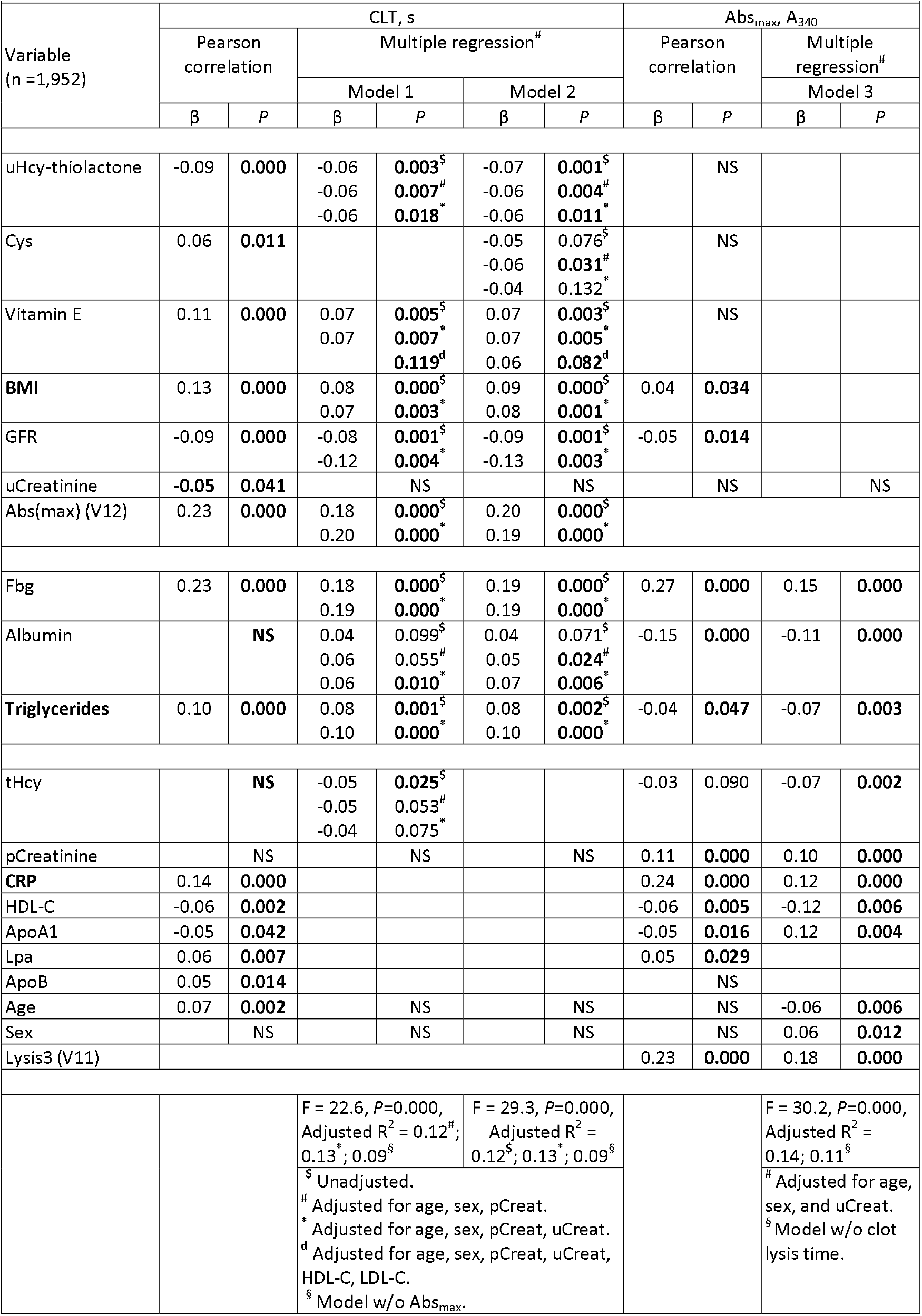
Determinants of plasma fibrin clot lysis time (CLT) and maximum absorbance (Abs_max_)

In multiple regression analysis, both fibrin CLT and Abs_max_ were associated with fibrinogen, albumin, and TG (**Table 2**). However, while fibrin CLT and Abs_max_ were positively associated with fibrinogen, CLT and Abs_max_ showed opposite associations with albumin and TG: positive for CLT and negative for Abs_max_.

Fibrin CLT and Abs_max_ showed disparate associations with other variables in multiple regression analysis. For example, fibrin CLT, but not Abs_max_, was associated with vitamin E, BMI, GFR, TG, and fibrinogen. On the other hand, Abs_max_, but not CLT, was associated with pCreatinine, CRP, HDL-C, and ApoA1 (**Table 2**).

Many associations found in bivariate analysis were not observed in multiple regression analysis. Specifically, although fibrin CLT was associated with uCreatinine, CRP, HDL-C, ApoA1, Lpa, ApoB, and age in bivariate analysis, these associations were not observed in multiple regression analysis (**Table 2**). Similarly, Abs_max_ was associated with BMI, GFR, and Lpa in bivariate analysis, but not in multiple regression analysis. Conversely, associations of Abs_max_ with tHcy, age, and sex found in multiple regression analysis, were not observed in bivariate analysis (**Table 2**).

The associations of uHcy-thiolactone, vitamin E, and albumin with fibrin CLT persisted in multiple regression models adjusted for age, sex, pCreatinine, and uCreatinine, as did the associations of BMI, GFR, and triglycerides with CLT (**Table 2**). In contrast, the association of tHcy or Cys with fibrin CLT lost significance in models adjusted for pCreat or uCreat, respectively. The association of vitamin E with fibrin CLT lost significance in models adjusted for HDL-C and LDL-C. Additional adjustments for CRP, ApoB, ApoA1, Lpa, tCys/tHcy had no effect on these associations.

In addition, baseline fibrin CLT was significantly longer in patients with hypercholesterolemia, diabetes, and obesity (*P* ≤ 0.003) but was unaffected by CVD status (**Table 3**). There was a tendency for longer CLT in patients with hypertension (P = 0.051) and previous acute myocardial infarction (AMI) (*P* = 0.066). In contrast, fibrin clot Abs_max_ was not affected by the disease status (**Table 3**).

**Table 3.**
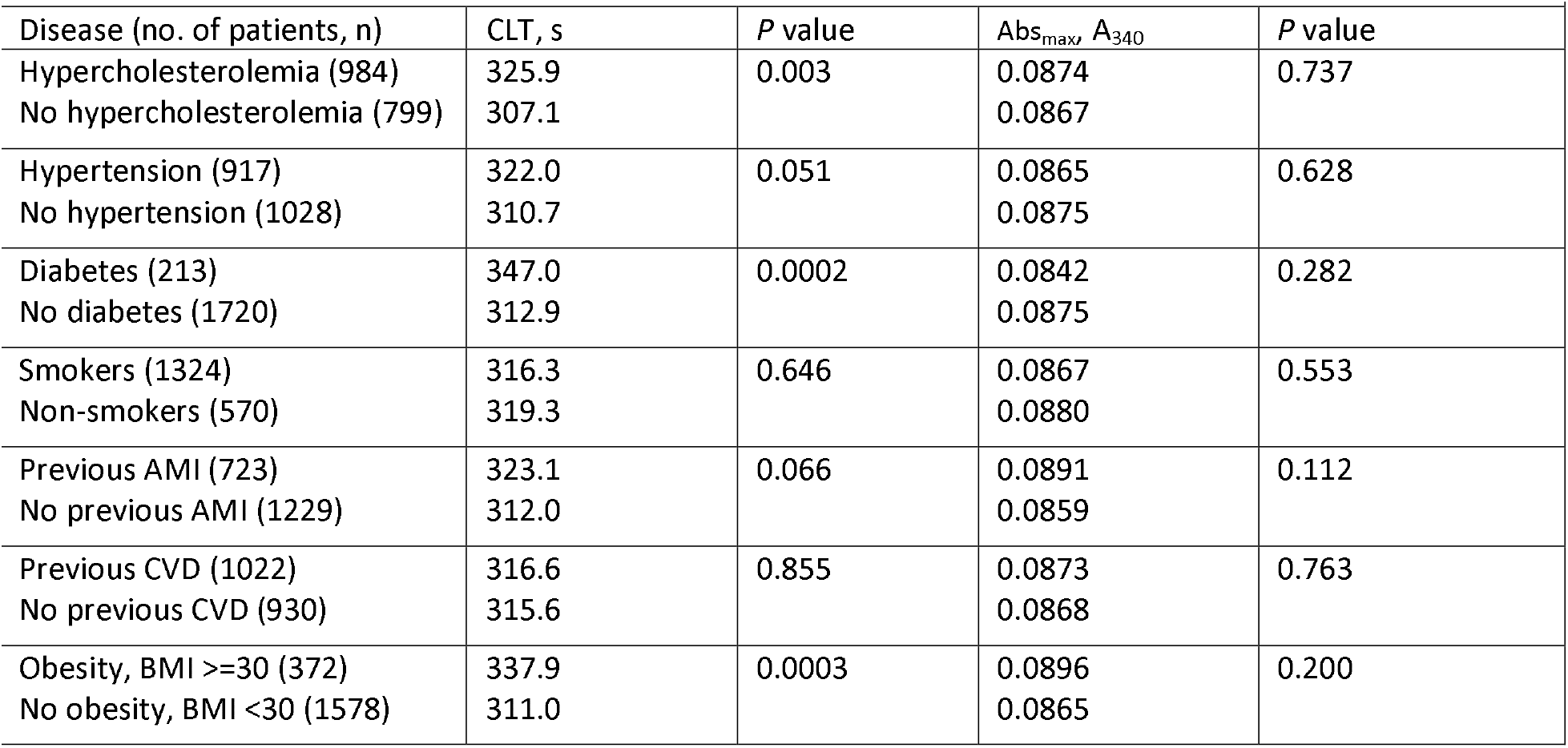
Fibrin clot lysis time (CLT) and maximum absorbance (Abs_max_) in CAD patients stratified by disease status at baseline

|  |  |  |  |  |
| --- | --- | --- | --- | --- |
| Disease (no. of patients, n) | CLT, s | *P* value | Abs<sub>max</sub> A<sub>340</sub> | *P* value |
| Hypercholesterolemia (984) | 325.9 | 0.003 | 0.0874 | 0.737 |
| No hypercholesterolemia (799) | 307.1 |  | 0.0867 |  |
| Hypertension (917) | 322.0 | 0.051 | 0.0865 | 0.628 |
| No hypertension (1028) | 310.7 |  | 0.0875 |  |
| Diabetes (213) | 347.0 | 0.0002 | 0.0842 | 0.282 |
| No diabetes (1720) | 312.9 |  | 0.0875 |  |
| Smokers (1324) | 316.3 | 0.646 | 0.0867 | 0.553 |
| Non-smokers (570) | 319.3 |  | 0.0880 |  |
| Previous AMI (723) | 323.1 | 0.066 | 0.0891 | 0.112 |
| No previous AMI (1229) | 312.0 |  | 0.0859 |  |
| Previous CVD (1022) | 316.6 | 0.855 | 0.0873 | 0.763 |
| No previous CVD (930) | 315.6 |  | 0.0868 |  |
| Obesity, BMI ≥30 (372) | 337.9 | 0.0003 | 0.0896 | 0.200 |
| No obesity, BMI <30 (1578) | 311.0 |  | 0.0865 |  |

### Impact of fibrin CLT and Abs_max_ on acute myocardial infarction and mortality

During the median follow-up time of 7 years, there were 8.0% (n = 160) acute myocardial infarctions (AMI) and mortality was 5.8% (n = 116), with a higher proportion of events in groups with a longer plasma fibrin CLT (>397.5 s) or higher Abs_max_ (>0.025) (**Table S4**). Kaplan-Meier analysis showed a worse survival free of AMI events among CAD patients with a longer fibrin CLT (>397.5 s) compared to patients with a shorter fibrin CLT (≤397.5) (*P* = 0.011) (**Figure 3A**). Survival free of mortality was also worse in CAD patients with a longer fibrin CLT (>532.5 s) compared to patients with a shorter fibrin CLT (≤532.5 s) (*P* = 0.002) (**Figure 3B**). Kaplan-Meier analysis also showed worse survival without an AMI (**Figure 3C**) or mortality (**Figure 3D**) among CAD patients with a higher Abs_max_ compared to those with lower Abs_max_. These differences started early in year 1 and progressively increased in years 2 - 7 during the follow-up.

**Figure 3.**
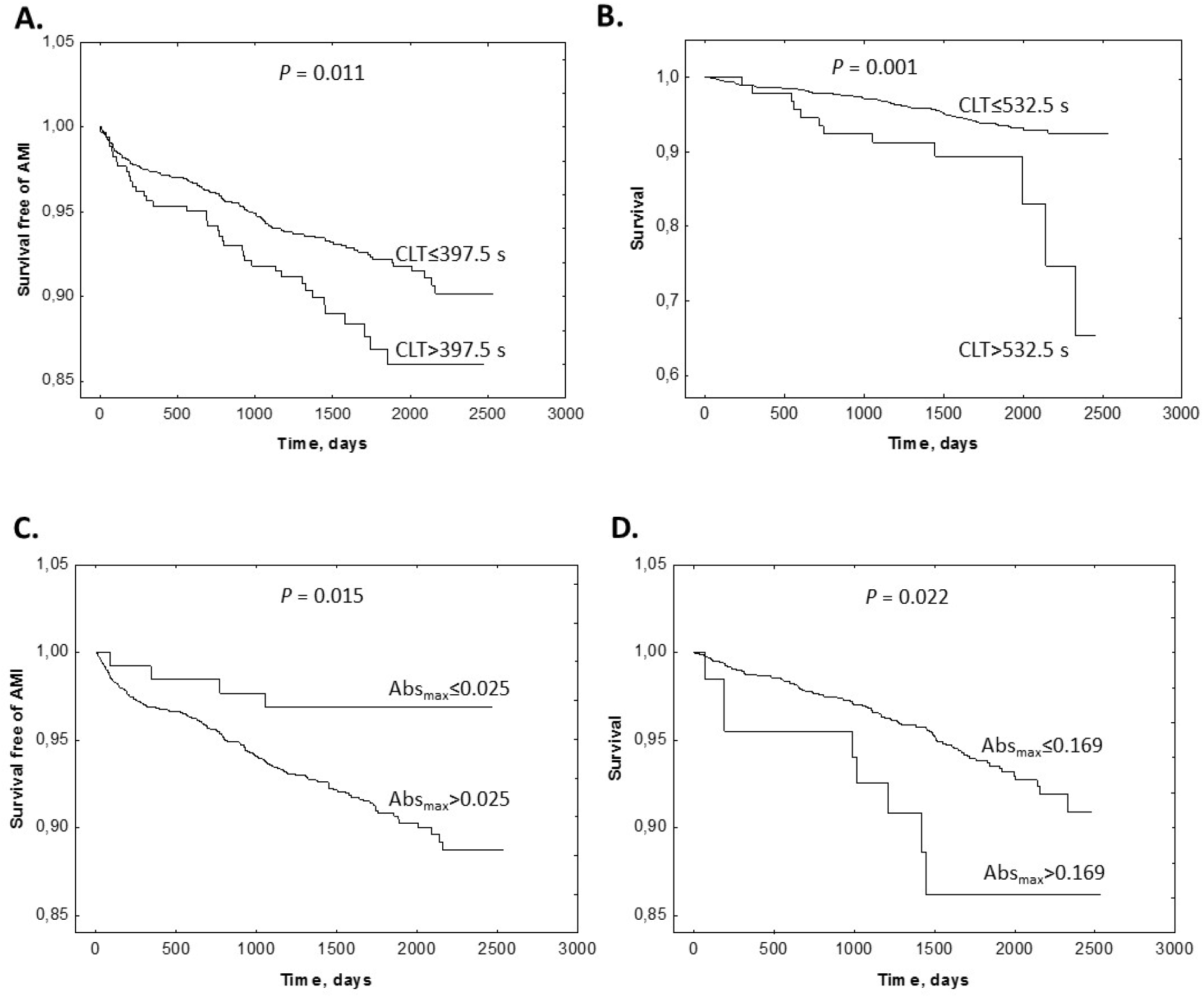
Kaplan-Meier analysis of outcome events according to CLT and Abs_max_ cutoffs. **A**. Survival free of AMI in CLT group 0 (CLT ≤ 397.5 s) and group 1 (CLT > 397.5) *vs*. time (days). **B**. Survival without mortality in CLT group 0 (CLT ≤ 532.5 s) and group 1 (CLT > 532.5 s). **C**. Survival free of AMI in Abs_max_ group 0 (Abs_max_ ≤ 0.025) and group 1 (A_max_ > 0.025) *vs*. time (days). **D**. Survival without mortality in Abs_max_ group 0 (Abs_max_ ≤ 0.169) and group 1 (Abs_max_ > 0.169).

Multivariable Cox proportional hazard regression analysis in a model including age and sex, showed that fibrin CLT (>cutoff value) was significantly associated with the incidence of AMI (HR 1.58, CI 1.10-2.28; *P* = 0.013) and mortality (HR 2.54, CI 1.40-4.63; *P* = 0.002) (Model 1, **Table 4**). The association between fibrin CLT and AMI remained significant after adjustment for vitamin E (HR 1.50, CI 1.04-2.18, *P* = 0.031; Model 2), glucose and BMI (HR 1.53, CI 1.07-2.21, *P* = 0.021; Model 3), LDL-C, HDL-C, TG, APOA1, APOB, and Lpa (HR 1.46, CI 1.01-2.11, *P* = 0.042; Model 4), uHcy-thiolactone, tHcy, and Cys (HR 1.57, CI 1.10-2.27, *P* = 0.014; Model 6), and uHcy-thiolactone, tHcy, Cys, uCreatinine, pCreatinine, and GFR (HR 1.53, CI 1.05-2.23, *P* = 0.025; Model 5). However, adjustments for smoking, diabetes, hypertension, extent of CAD at angiography, LVEF, heart failure, previous peripheral artery disease, AMI, stroke, and coronary artery bypass attenuated the association between CLT and AMI (HR 1.43, CI 0.99-2.06, *P* = 0.058; Model 7, Table 5), as did adjustments for fibrinogen and CRP (HR 1.41, CI 0.96-2.08, P=0.081; Model 8, **Table 4**).

**Table 4.** Impact of plasma fibrin clot lysis time (CLT) and maximum absorbance (Abs_max_) on HR (95% CI) for outcomes - Cox regression.*

|  |  |  |  |  |  |  |  |  |
| --- | --- | --- | --- | --- | --- | --- | --- | --- |
| CLT, 0,1 | CLT (n = 1,952) |  |  |  | Abs<sub>max</sub> (n = 1,952) |  |  |  |
|  | Mortality (n = 116, 5.8%) |  | AMI (n = 160, 8.0%) |  | Mortality (n = 116, 5.8%) |  | AMI (n = 160, 8.0%) |  |
|  | HR (95% CI) | P value | HR (95% CI) | P value | HR (95% CI) | P value | HR (95% CI) | P value |
| Model 1<sup>a</sup> | 2.54 (1.40-4.63) | 0.002 | 1.58 (1.10-2.28) | 0.013 | 2.39 (1.17-4.92) | 0.017 | 3.22 (1.19-8.69) | 0.021 |
| Model 2<sup>b</sup> | 2.55 (1.36-4.77) | 0.003 | 1.50 (1.04-2.18) | 0.031 | 2.31 (1.12-4.74) | 0.023 | 2.98 (1.10-8.07) | 0.031 |
| Model 3<sup>c</sup> | 2.67 (1.46-4.86) | 0.001 | 1.53 (1.06-2.21) | 0.021 | 2.44 (1.19-5.01) | 0.015 | 3.12 (1.15-8.44) | 0.025 |
| Model 4<sup>d</sup> | 2.42 (1.32-4.44) | 0.004 | 1.46 (1.01-2.11) | 0.042 | 2.10 (1.01-4.36) | 0.047 | 3.24 (1.19-8.77) | 0.021 |
| Model 5<sup>e,#,§</sup> | 2.67 (1.42-5.01) | 0.002 | 1.53 (1.05-2.23) | 0.025 | 2.11 (0.97-4.58) | 0.059 | 2.90 (1.07-7.84) | 0.036 |
| Model 6<sup>f,#</sup> | 2.70 (1.48-4.93) | 0.001 | 1.57 (1.09-2.26) | 0.014 | 2.15 (1.05-4.43) | 0.037 | 3.18 (1.18-8.60) | 0.022 |
| Model 7<sup>g</sup> | 2.60 (1.42-4.77) | 0.002 | 1.43 (0.99-2.06) | 0.058 | 2.02 (0.93-4.38) | 0.073 | 3.31 (1.22-8.97) | 0.019 |
| Model 8<sup>h</sup> | 2.24 (1.13-4.46) | 0.021 | 1.41 (0.96-2.08) | 0.081 | 1.69 (0.75-3.77) | 0.202 | 3.13 (1.16-8.47) | 0.025 |
| *HR, hazard ratio; CI, confidence interval. CLT was included as a categorical term at cut-off values of 532.5 s for mortality and 397.5 s for AMI: 0, CLT < cutoff; 1, CLT ≥ cutoff. Abs<sub>max</sub> was included as a categorical term at cut-off values of 0.169 A<sub>340</sub> for mortality, 0.025 A<sub>340</sub> for AMI: 0, Abs<sub>max</sub> < cutoff; 1, Abs<sub>max</sub> ≥ cutoff. Sex: 0, female; 1, male. Other variables were continuous.<br><sup>a</sup> Adjusted for age and sex, <sup>b</sup> adjusted as in Model 1 plus vitamin E; <sup>c</sup>, adjusted as in Model 1 plus glucose and BMI; <sup>d</sup>, adjusted as in Model 1 plus LDL-C, HDL-C, TG, APOA1, APOB, and Lpa. <sup>e</sup>, adjusted as in Model 1 plus uHcy-thiolactone, tHcy, Cys, uCreatinine, pCreatinine, and GFR; <sup>f</sup> adjusted as in Model 1 plus uHcy-thiolactone, tHcy, Cys; <sup>g</sup>, adjusted as in Model 1 plus smoking, diabetes, hypertension, extent of CAD at angiography, LVEF, heart failure, previous peripheral artery disease, AMI, stroke, and coronary artery bypass; <sup>h</sup>, adjusted as in Model 1 plus fibrinogen and CRP.<br><sup>§</sup> CLT: uHcy-thiolactone was significantly associated with AMI in model 5 (HR per 100 nM increase in uHcy-thiolactone was 1.12, CI 1.01-1.20; P = 0.031).<br><sup>#</sup> CLT and Abs<sub>max</sub>: Cys was significantly associated with AMI. Model 5: HR per 100 μM increase in Cys was 1.7, CI 1.2-10.1; P = 0.009. Model 6: HR per 100 μM increase in Cys was 1.6, CI 1.2-11.0; P = 0.009. tHcy was not associated with AMI in Models 7 and 8: HR per 10 μM increase in tHcy were 1.1 and 1.2, CI 0.99-1.04; P = 0.182 and 0.289. |  |  |  |  |  |  |  |  |

**Table 5.** Plasma fibrin clot lysis time (CLT) and maximum absorbance (Abs_max_) at the end of study according to folic acid and B-vitamin supplementation status

|  |  |  |  |  |  |  |  |
| --- | --- | --- | --- | --- | --- | --- | --- |
| Variable | Treatment group<sup>a</sup> |  |  |  |  |  |  |
|  | Folic acid + vitamins B<sub>12</sub>, B<sub>6</sub> |  | Folic acid + vitamin B<sub>12</sub> |  | Vitamin B<sub>6</sub> |  | Placebo |
|  | Mean ± SD | *P* value vs. placebo | Mean ± SD | *P* value vs. placebo | Mean ± SD | *P* value vs. placebo | Mean ± SD |
| CLT, s | 322 ± 101 | 0.369 | 333 ± 115 | 0.638 | 327 ± 89 | 0.476 | 329 ± 91 |
| Abs<sub>max</sub>, A<sub>340</sub> | 0.095 ± 0.033 | 0.859 | 0.090 ± 0.029 | 0.240 | 0.104 ± 0.038 | 0.318 | 0.097 ± 0.036 |
| Plasma Cys, μM | 295 ± 31 | 0.516 | 292 ± 35 | 0.804 | 286 ± 36 | 0.481 | 291 ± 33 |
| Plasma tHcy, μM | 7.52 ± 1.48 | **1.E-10** | 7.61 ± 1.68 | **1.E-09** | 10.72 ± 3.54 | 0.739 | 10.51 ± 2.58 |
| uHcy-thiolactone, nM | 59.5 ± 58.6 | 0.371* | 91.2 ± 106.0 | 0.571* | 83.3 ± 133.6 | 0.775* | 61.3 ± 49.8 |
| Folic acid | 63.2 ± 32.5 | **4.E-14** | 71.0 ± 32.2 | **2.E-17** | 11.3 ± 8.2 | 0.063 | 16.7 ± 18.6 |
| Vitamin B<sub>12</sub> | 611 ± 183 | **2.E-6** | 617 ± 218 | **2.E-5** | 365 ± 129 | 0.260 | 409 ± 246 |
| Vitamin B<sub>6</sub> | 307 ± 182 | **2.E-16** | 40.2 ± 19.6 | 0.157 | 275 ± 167 | **2.E-15** | 48.8 ± 37.7 |
|  | <sup>a</sup> n = 52 for each group. CLT and Abs<sub>max</sub> data were obtained for n = 46-50 patients in each group. * *P* values for log-transformed data. |  |  |  |  |  |  |

In Cox regression models that included CLT and sulfur-containing metabolites (Models 5 and 6), Cys and uHcy-thiolactone, but not tHcy, were significant predictors of AMI (**Table 4**).

Cox regression analysis in a model including age and sex, showed that fibrin Abs_max_ (>cutoff value) was significantly associated with the incidence of AMI (HR 3.22, CI 1.19-8.69; *P* = 0.021) and mortality (HR 2.39, CI 1.17-4.92; *P* = 0.017) (Model 1, **Table 4**). The association between fibrin CLT and AMI remained significant after adjustment for vitamin E (HR 2.98, CI 1.10-8.07, *P* = 0.031; Model 2), glucose and BMI (HR 3.12, CI 1.15-8.44, *P* = 0.025; Model 3), LDL-C, HDL-C, TG, APOA1, APOB, and Lpa (HR 3.24, CI 1.19-8.77, *P* = 0.021; Model 4), uHcy-thiolactone, tHcy and Cys (HR 3.18, CI 1.18-8.60, *P* = 0.022; Model 6), and uHcy-thiolactone, tHcy, Cys, uCreatinine, pCreatinine, and GFR (HR 2.90, CI 1.07-7.84, *P* = 0.036; Model 6). Adjustments for smoking, diabetes, hypertension, extent of CAD at angiography, LVEF, heart failure, previous peripheral artery disease, AMI, stroke and coronary artery bypass (HR 3.31, CI 1.22-8.97, *P* = 0.019; Model 7, **Table 4**) and for fibrinogen and CRP (HR 3.13, CI 1.16-8.47, *P* = 0.025; Model 8, **Table 4**).

### Supplementation with folic acid, vitamin B_12_ and/or vitamin B_6_ did not affect fibrin CLT and Abs_max_

As previously reported [11], supplementation with folic acid, vitamin B_12_ and/or vitamin B_6_ resulted in significant increases in plasma levels of these vitamins and a significant decrease in plasma tHcy at the end of study (**Table 5**). However, we found that the supplementation with any combination of folic acid and B vitamins did not affect fibrin CLT or Abs_max_ (**Table 5**). This finding indicates that the reduction in plasma tHcy does not affect fibrin clot structure/function and is consistent with the absence of correlation between fibrin CLT or Abs_max_ and plasma tHcy at baseline (**Figure 1C**). We also found that uHcy-thiolactone and plasma Cys, which were correlated with fibrin CLT at baseline (**Figure 1A, B**), were not affected by the supplementation with any combination of folic acid and B vitamins (**Table 5**).

## Discussion

We found that at baseline, (*i*) uHcy-thiolactone and plasma Cys were significantly associated with CLT, while plasma Hcy was a significantly determinant of Abs_max_, independent of fibrinogen, triglycerides, vitamin E, GFR, independently of fibrinogen, triglycerides, vitamin E, creatinine, CRP, HDL-C, ApoA1, GFR, BMI, age, and sex; (*ii*) Kaplan-Meier analyses showed worse survival and worse survival free of AMI in CAD patients with longer baseline CLT and higher Abs_max_; (*iii*) in Cox regression analyses, longer baseline CLT and higher Abs_max_ predicted future AMI events and mortality; (*iv*) in Cox regression models with CLT, Cys and uHcy-thiolactone predicted AMI, while Cys predicted AMI in Cox models with Abs_max_. (*v*) B-vitamin/folic acid therapy did not affect CLT and Abs_max_.

Accumulating evidence suggests that dysregulation of sulfur amino acid metabolism and impaired fibrin clot properties are associated with CVD. In the present work we examined Hcy-thiolactone, Hcy, and Cys as determinants of fibrin CLT and Abs_max_. We found that CLT and Abs_max_ were significantly correlated with each other (β = 0.23, *P* = 0.000) in a cohort of CAD patients, consistent with previous findings in studies with healthy probands [2] and patients with diabetes [13][14].

Notably, in multiple regression analysis, we found that uHcy-thiolactone was a negative predictor (β=-0.07, P=0.001) while Cys was a positive predictor (β=0.06, P=0.031) of CLT. Association of uHcy-thiolactone with CLT was independent of vitamin E, which we identified as another new predictor of CLT (β=0.07, P=0.003) (**Table 2**). The association of CLT with vitamin E is consistent with previous findings showing that vitamin E is an inhibitor of plasmin-mediated fibrinolysis [20].

The association of CLT with uHcy-thiolactone was also independent of BMI, GFR, CRP, fibrinogen, and TG, known determinants of CLT. Importantly, uHcy-thiolactone remained a significant predictor of CLT after adjustments for age, sex, tHcy, pCreatinine, and uCreatinine (**Table 2**) as well as ApoB, Lpa, HDL-C, ApoA1 (not shown). However, the association between plasma Cys and CLT was attenuated by the adjustment for uCreatinine. We found that tHcy was significantly associated with CLT only in an unadjusted model but not in models adjusted for age, sex, pCreatinine, and uCreatinine (**Table 2**). Another study with large cohorts of thrombosis patients (n = 770) and healthy controls (n = 743) also reported no association of CLT with tHcy [21].

Other studies reported that CLT was associated with tHcy [22], CRP [23], Lpa [24], HDL-C and ApoA1 [25]. However, these were small studies (about 100 subjects) and different adjustments for possible confounders (or a lack thereof), compared with the size of a cohort (n=1,952) and adjustments used in the present study (**Table 2**). Further, although we found that CRP, HDL-C, ApoA1, Lpa, and ApoB, but not tHcy, were significantly associated with CLT in a univariate analysis, these associations were absent in multivariate regression analysis (**Table 2**).

Notably, in multiple regression analyses, we found a significant negative association of Abs_max_ with Hcy (β=-0.07, P=0.002) that has not been reported before. The association between tHcy and Abs_max_ was not affected by other variables associated with Abs_max_ in our cohort: albumin, TG, HDL-C; ApoA1, fibrinogen, pCreatinine, CRP, age, and sex. Adjustments for uHcy-thiolactone, Cys, BMI, GFR, uCreatinine,Lpa, LDL-C, and vitamin E did not affect these associations. This finding suggests that Hcy may affect the structure of the plasma fibrin clot without affecting clot’s function (see below).

The disparate effects of uHcy-thiolactone and tHcy on CLT and Abs_max_ suggest that each metabolite can affect clot properties *via* different metabolite-specific mechanisms. Some metabolites may affect clot structure while others might affect function. For example, Hcy-thiolactone has the ability to modify protein lysine residues in a process called *N*-hcomocystinylation, which leads to protein damage [5]. The accumulation of Hcy-thiolactone in the blood can be harmful because it will lead to increased levels of *N*-Hcy-protein [26]. More efficient urinary clearance of Hcy-thiolactone [8] will reduce its blood concentration and *N*-Hcy-protein levels, including *N*-Hcy-fibrinogen and *N*-Hcy-albumin [27]. *N*-Hcy-fibrinogen is prothrombotic because it forms fibrin clots that are more resistant to lysis by plasmin [28]. Hcy and Cys have the ability to bind to proteins via disulfide bonds [5]. Indeed, most of Hcy and Cys present in human plasma is carried on plasma proteins. Specifically, *N*-Hcy-fibrinogen and *N*-Hcy-albumin as well as S-Hcy-albumin and S-Cys-albumin have been identified in human plasma [29].

As fibrin clot contains other protein components, in addition to fibrinogen, variations in the levels of these proteins, their modified forms (*e*.*g*., *N*-Hcy-fibrinogen, *N*-Hcy-albumin, *S*-Hcy-albumin), as well as variations in sulfur-containing metabolites (*e*.*g*., Hcy-thiolactone, Hcy, Cys), will result in fibrin clots of different composition, structure, and susceptibility to lysis. These factors most likely account for the associations of the sulfur-containing metabolites as well as fibrinogen and albumin with measures of fibrin clot properties such as CLT and Abs_max_.

In this context, our findings suggest that attenuated uHcy-thiolactone excretion can be harmful, because it would elevate plasma Hcy-thiolactone and *N*-Hcy-fibrinogen, which would generate fibrin clots resistant to lysis, reflected in longer CLT. On the other hand, elevation of plasma Cys, which would elevate plasma *S*-Cys-albumin and lead to the incorporation of more *S*-Cys-albumin into fibrin clot structure, can be beneficial, because it is associated with increased susceptibility of fibrin clots to lysis, reflected in shorter CLT. Our findings also suggest that elevated plasma tHcy, which would lead to the incorporation of more S-Hcy-protein into fibrin clot structure, is associated with reduced Abs_max_, which suggests a less compact fibrin clot structure. However, a presumably less compact fibrin clot structure in this case apparently didn’t improve fibrin clot susceptibility to lysis, as reflected by the absence of any significant association of tHcy with CLT.

This interpretation is consistent with our present findings showing that CLT and Abs_max_ predict AMI and mortality, independent of other risk factors. In the present study we also found that plasma Cys is a new independent determinant of CLT and a predictor of AMI and mortality. Further, we have identified uHcy-thiolactone as a new independent determinant of CLT (**Table 2**) and confirmed (**Table 4**, Model 5) our previous finding that uHcy-thiolactone is a predictor of AMI [6].

In the present study, we found that longer CLT and higher Abs_max_ predicted AMI (**Table 4**). Previous studies also have shown a positive association between plasma fibrin clot properties and outcomes. For example, a case-control study with AMI patients (n = 800) and controls (n = 1,123) showed that long CLT and high Abs_max_ (>90th percentile in controls) were associated with an increased risk of AMI, 2.62- and 1.66-fold, respectively [30].

Another study with 300 patients hospitalized with acute coronary syndrome found that long CLT (>77^th^ percentile) was associated with a 2.52-fold increased risk of major adverse cardiovascular events (a composite of CV death, nonfatal AMI, and stroke) at a 12-month follow-up [31]. CLT and Abs_max_ predicted AMI and mortality in a large PLATO study involving patients with acute coronary syndrome (n = 4,354; 138 CV death events, 145 all-cause death, 183 AMI, 41 stroke, 256 major bleeding, 96 non-coronary artery bypass graft-related major bleeding) [32] and patients with diabetes (n = 974; 48 CV death events, 72 AMI, 67 major bleeding, 21 non-coronary artery bypass graft-related major bleeding) [14]. However, these prospective studies involved a short 1-year follow-up and, except for the PLATO study, a limited rate of events, compared to the present study of 1,952 CAD patients, which found that longer CLT and higher Abs_max_ predicted AMI (n = 160 events) and mortality (n = 116) during a 7-year follow-up (**Table 4**). In a relatively small study with CAD patients (n = 786; composite of nonfatal AMI, ischemic stroke, and cardiovascular death, n = 70), CLT or Abs_max_ did not predict vascular events after a 3-year follow-up [33], possibly due to its smaller size.

Previous studies have suggested that the failure of B-vitamin to attenuate Hcy-thiolactone levels [6], anti-*N*-Hcy-protein autoantibodies [34], and inflammation [34, 35] could account for the lack of efficacy of tHcy-lowering B-vitamin intervention trials in alleviating AMI [36]. Our present findings provide an additional explanation that can account for the lack of efficacy of the B-vitamin therapy: B-vitamin therapy was ineffective because it did not improve fibrin clot properties (CLT and Abs_max_), an important pro-atherogenic factor.

It is generally accepted in the field that Abs_max_ reflects the clot structure while CLT is a functional property that reflects clot’s susceptibility to lysis. High Abs_max_ indicates a more compact structure of dense thin fibers characterized by low permeability and low susceptibility to lysis, while low Abs_max_ indicates a less compact structure of loose thicker fibers characterized by high permeability and high susceptibility to lysis [37]. These structures have been identified by electron microscopy. Thus, one can expect that factors affecting Abs_max_ should also similarly affect CLT and vice versa. However, we found that factors affecting Abs_max_ did not affect CLT and factors affecting CLT did not affect Abs_max_ (**Table 2**). Specifically, in multiple regression analysis we identified variables that were associated with (*i*) only plasma fibrin CLT (uHcy-thiolactone, Cys, vitamin E, BMI, and GFR) and (*ii*) only with plasma fibrin clot Abs_max_ (tHcy, pCreatinine, CRP, HDL-C, APOA1, age, and sex). Only three variables were associated with both plasma fibrin CLT and Abs_max_ (fibrinogen, albumin, and triglycerides) (**Table 2**).

The lack of congruence between associations of some variables with CLT and Abs_max_ suggests that variables that affect only CLT may act on a functional component of the fibrinolysis cascade without affecting Abs_max_, *i*.*e*., clot structure. A case in point is vitamin E, which affected CLT, but not Abs_max_, and is known to inhibit plasmin-mediated fibrinolysis [20]. On the other hand, variables that affect only Abs_max_ apparently involve changes in the clot structure that have no functional consequence on CLT. Only a few variables were correlated with both structure and function of the fibrin clot (Table 2).

### Strength and limitations

The present study is the first to evaluate sulfur-containing compounds as determinants of fibrin clot properties in relation to future AMI events and mortality in CAD patients. Major strengths of the present study are the size (n = 1,952), the prospective design, an extended 7-year-long follow-up, and an extensive information regarding baseline clinical/biochemical characteristics. Further, the high E-values for HR and lower CI suggest that our findings are robust to the presence of unmeasured confounders. Thus, these findings are likely to be reproducible in other populations. As our study was limited to a white elderly population with CAD, our findings remain to be confirmed in populations in other age groups and ethnic backgrounds.

## Conclusions

uHcy-thiolactone and plasma Cys are novel determinants of CLT. Plasma Hcy is a determinant of Abs_max_ but not of CLT. CLT and Abs_max_ predict future AMI events and mortality in CAD patients but were not affected by B-vitamin/folate supplementation. Our findings also suggest that targeting sulfur-containing metabolites other than tHcy might be a useful therapeutic strategy to mitigate prothrombotic phenotypes that increase a risk of AMI and mortality.

## Data Availability

All data produced in the present work are contained in the manuscript

## Supplementary material

Supplementary material is available online.

## Acknowledgements

We thank Peter Grant for kindly providing software for the analysis of plasma fibrin clot formation and lysis. This work was supported in part by grants from the National Science Center, Poland: 2016/23/B/NZ5/00573, 2018/29/B/NZ4/00771, 2019/33/B/NZ4/01760.

## Author contribution

MS, PS measured kinetics of fibrin clot formation and lysis, calculated fibrin clot lysis time and maximum turbidity; JP-K analyzed the data; ON provided citrated pasma samples from CAD patients and the database for the WENBIT cohort; HJ conceived the idea for the project, designed the research, analyzed the data, wrote the manuscript, and had a primary responsibility for the final content; and all authors have read and approved the final manuscript.

