## Supplemental material for "Homocysteine thiolactone contributes to a prognostic value of fibrin clot structure/function in coronary artery disease patients"

**Supplementary material**

**Figure S1**

**Table S1**

**Table S2**

**Table S3**

**Table S4**

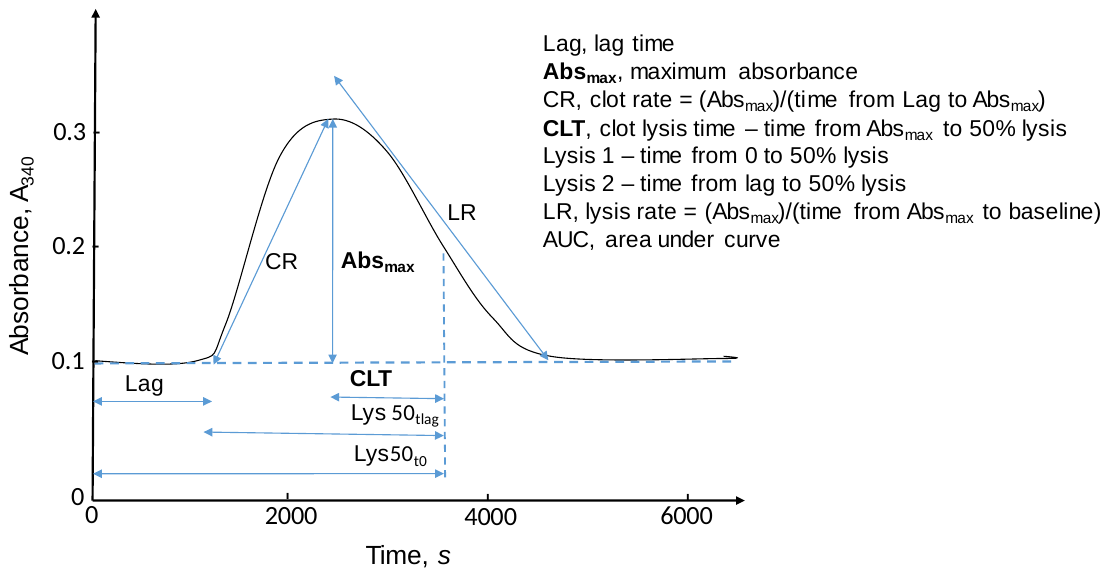

**Figure S1** Illustration of clotting and lysis variables. Variables examined in a greater detail in the present study, CLT and Abs_max_, are highlighted in bold.

| **Table S1** Pearson correlation coefficients for relationships between turbidimetric clotting and lysis variables in the WENBIT cohort of CAD patients (n = 1,983) ^a^ | | | | | | | |
| --- | --- | --- | --- | --- | --- | --- | --- |
|  | MaxAbs, **Abs_max_**^b^ | Clot Rate | Lys50_t0_ | Lys50_tlag_ | Lysis50_MA_, **CLT** ^b^ | Lysis Rate | LysisArea  AUC |
| Lag | **-0.61** | **-0.54** | **0.46** | **-0.20** | **-0.08** (*P*=0.001) | **0.41** | **0.05** (*P*=0.028) |
| AbsMax,  **Abs_max_**^b^ |  | **0.87** | **-0.17** | **0.26** | **0.23** | **-0.69** | **0.27** |
| Clot Rate |  |  | **-0.41** | **-0.07** (*P*=0.002) | 0.00 (*P*=0.899) | **-0.67** | **0.16** |
| Lys50_t0_ |  |  |  | **0.77** | **0.73** | **0.28** | **0.24** |
| Lys50_tlag_ |  |  |  |  | **0.86** | 0.02  (*P*=0.407) | **0.23** |
| Lysis50_MA_, **CLT**^b^ |  |  |  |  |  | **0.05** (*P*=0.028) | **0.29** |
| Lysis Rate |  |  |  |  |  |  | **-0.11** |
| ^a^ Nomenclature after Carter *et al*., *Arterioscler Thromb Vasc Biol* 2007; 27:2783-2789.  *P*<0.000, except when indicated otherwise. The clotting and lysis variables are illustrated in **Figure S1**.  ^b^ Terms **Abs_max_** and **CLT** referring to terms MaxAbs and Lysis50_MA_, respectively, of Carter *el al*. ATVB 2007, have been use in the present study. | | | | | | | |

| **Table S2** Biochemical characteristics across quartiles of plasma fibrin clot lysis time (CLT) in CAD patients | | | | | |
| --- | --- | --- | --- | --- | --- |
| Variable (n=1,952) | CLT quartiles | | | |  |
|  | Q1 (<240 s)  N=464 | Q2 (240-300 s)  N=638 | Q3 (>300-360 s)  N=391 | Q4 (>360 s)  N=459 | ANOVA  *P*-value |
| uHcy-thiolactone, nM | 46.3 (18.8-117.4) | 50.1 (23.5-107.9) | 46.9 (22.6-88.8) | 39.9 (20.5-85.1) | 0.001 |
| tHcy, μM | 10.6 (8.7-12.9) | 10.8 (9.1-12.5) | 10.5 (8.8-12.7) | 10.7 (8.8-13.0) | 0.619 |
| Cys, μM | 287 (265-310) | 289 (267-310) | 292 (271-316) | 295 (272-320) | 0.006 |
| uCreatinine, mM | 8.9 (6.1-12.0) | 8.8 (5.9-11.9) | 8.1 (5.6-11.4) | 8.3 (5.7-10.8) | 0.010 |
| Vitamin E | 28.9 (22-33) | 30.1 (26-34) | 30.9 (27-35) | 31.2 (27-36) | 0.000 |
| pCreatinine, μM | 72 (65-83) | 76 (66-85) | 76 (66-85) | 75 (66-85) | 0.259 |
| Albumin, g/L | 43 (42-45) | 43 (42-45) | 43 (41-45) | 43 (41-45) | 0.021 |
| Fibrinogen, g/L | 3.3 (2.9-3.7) | 3.50 (3.1-3.9) | 3.6 (3.2-4.2) | 3.9 (3.4-4.3) | 0.000 |
| CRP, mg/L | 1.3 (0.7-2.6) | 1.4 (0.8-3.1) | 2.2 (1.0-3.8) | 2.4 (1.1-5.4) | 0.000 |
| TG, mM | 1.26 (0.97-1.85) | 1.50 (1.10-2.09) | 1.51 (1.08-2.16) | 1.59 (1.11-2.27) | 0.002 |
| LDL-C | 2.9 (2.3-3.7) | 3.0 (2.4-3.7) | 3.0 (2.5-3.7) | 3.0 (2.4-3.9) | 0.329 |
| HDL-C, mM | 1.3 (1.1-1.6) | 1.2 (1.0-1.5) | 1.2 (1.0-1.5) | 1.2 (1.0-1.5) | 0.004 |
| APOA1, mg/dL | 1.3 (1.2-1.5) | 1.3 (1.2-1.5) | 1.3 (1.1-1.5) | 1.3 (1.1-1.5) | 0.411 |
| APOB, mg/dL | 0.85 (0.71-1.04) | 0.88 (0.75-1.05) | 0.89 (0.76-1.07) | 0.91 (0.76-1.10) | 0.003 |
| Lpa, mg/dL | 0.25 (0.13-0.52) | 0.27 (0.12-0.53) | 0.32 (0.16-0.62) | 0.34 (0.17-0.61) | 0.004 |
| BMI, kg/m^2^ | 25 (23-28) | 26 (23-28) | 26 (24-28) | 27 (24-30) | 0.000 |
| GFR, mL/min/1.73 m^2^ | 92 (80-101) | 91 (80-99) | 90 (78-98) | 87 (76-97) | 0.000 |
| Age, years | 61 (53-70) | 61 (55-70) | 62 (55-70) | 63 (56-71) | 0.000 |
| Absorption max, A_340_ | 0.059 (0.041-0.080) | 0.088 (0.068-0.106) | 0.099 (0.074-0.121) | 0.106 (0.065-0.140) | 0.000 |
|  | Values are medians (interquartile range) | | | | |

| **Table S3** Biochemical characteristics across quartiles of fibrin clot maximum absorbance (Abs_max_) in CAD patients | | | | | |
| --- | --- | --- | --- | --- | --- |
| Variable (n=1,952) | Abs_max_ quartiles | | | |  |
|  | Q1 (A_340_ < 0.057)  N=498 | Q2 (A_340_ 0.057-0.087)  N=491 | Q3 (A_340_ > 0.087-0.114)  N=486 | Q4 (A_340_ > 0.114-0.362)  N=459 | ANOVA  *P*-value |
| uHTL, nM | 42.1 (20.0-100.5) | 46.2 (20.0-105.7) | 45.0 (20.9-94.5) | 46.2 (22.5-94.9) | 0.824 |
| tHcy, μM | 10.8 (9.0-13.2) | 10.6 (8.7-12.6) | 10.4 (8.8-12.6) | 10.7 (8.9-12.8) | 0.014 |
| Cys, μM | 292 (268-315) | 288 (267-309) | 290 (269-315) | 294 (270-320) | 0.015 |
| uCreatinine, mM | 8.5 (5.7-11.4) | 8.9 (6.1-11.8) | 8.6 (6.1-11.3) | 8.2 (5.6-11.7) | 0.522 |
| Vitamin E | 31 (26-35) | 30 (26-35) | 30 (26-35) | 30 (26-35) | 0.967 |
| pCreatinine, μM | 73 (66-84) | 74 (65-83) | 75 (65-84) | 77 (67-86) | 0.009 |
| Albumin, g/L | 43 (42-45) | 43 (42-45) | 43 (42-45) | 43 (41-44) | 0.000 |
| Fibrinogen, g/L | 3.5 (3.0-4.0) | 3.4(3.1-3.9) | 3.5 (3.1-4.0) | 3.9 (3.4-4.4) | 0.000 |
| CRP, mg/L | 1.5 (0.8-3.0) | 1.4 (0.7-3.1) | 1.6 (0.8-3.8) | 2.4 (1.3-5.2) | 0.000 |
| TG, mM | 1.46 (1.05-2.13) | 1.48 (1.04-2.13) | 1.53 (1.11-2.12) | 1.42 (1.04-2.06) | 0.322 |
| LDL-C | 3.0 (2.4-3.8) | 3.0 (2.4-3.8) | 3.0 (2.4-3.8) | 2.9 (2.4-3.7) | 0.704 |
| HDL-C, mM | 1.2 (1.0-1.5) | 1.3 (1.0-1.5) | 1.2 (1.0-1.5) | 1.2 (1.0-1.4) | 0.304 |
| APOA1, mg/dL | 1.3 (1.2-1.5) | 1.3 (1.2-1.5) | 1.3 (1.1-1.5) | 1.3 (1.2-1.5) | 0.526 |
| APOB, mg/dL | 0.89 (0.74-1.08) | 0.87 (0.75-1.06) | 0.89 (0.75-1.06) | 0.88 (0.75-1.05) | 0.832 |
| Lpa, mg/dL | 0.26 (0.13-0.54) | 0.28 (0.13-0.56) | 0.29 (0.15-0.58) | 0.34 (0.16-0.61) | 0.082 |
| BMI, kg/m^2^ | 26 (23-28) | 26 (23-28) | 26 (24-29) | 26 (24-29) | 0.082 |
| GFR, mL/min/1.73 m^2^ | 90 (76-99) | 91 (80-100) | 90 (81-99) | 88 (77-97) | 0.002 |
| Age, years | 63 (55-71) | 61 (54-70) | 61 (55-69) | 62 (56-70) | 0.043 |
| Clot lysis time (CLT), s | 255 (195-345) | 270 (225-315) | 285 (255-345) | 345 (300-420) | 0.000 |
|  | Values are medians (interquartile range). | | | | |

| **Table S4** Groups of CAD patients stratified by outcome and cutoff values of  clot lysis time (CLT) or maximum absorbance (Abs_max_) | | | | | | | |
| --- | --- | --- | --- | --- | --- | --- | --- |
| Outcome group |  | Clot lysis time (CLT) | | | Maximum absorbance (Abs_max_) | | |
|  | No. of patients (%) | CLT cutoff value | No. of patients ≤cutoff (%) | No. of patients >cutoff (%) | CLT cutoff value | No. of patients ≤cutoff (%) | No. of patients >cutoff (%) |
| AMI | 2006 | 397.5 | 1637 | 345 | 0.025 | 131 | 1852 |
| 0 ^a^ | 1846 (92.0) |  | 1516 (92.6) | 306 (88.7) |  | 127 (96.9) | 1696 (91.6) |
| 1 ^a^ | 160 (8.0) |  | 121 (**7.4**) | 39 (**11.3**) |  | 4 (**3.1**) | 156 (**8.4**) |
| Mortality | 2006 | 532.5 | 1890 | 92 | 0.169 | 1916 | 67 |
| 0 ^a^ | 1890 (94.2) |  | 1786 (94.5) | 80 (87.0) |  | 1808 (94.4) | 59 (88.1) |
| 1 ^a^ | 116 (5.8) |  | 104 (**5.5**) | 12 (**13.0**) |  | 108 (**5.6**) | 8 (**11.9**) |
| ^a^ 0, group of patients without AMI or mortality events. 1, group of patients suffering from AMI or mortality. | | | | | | | |
